# Identification of an intrusive-hypervigilant phenotype of posttraumatic stress symptoms with unique stress peptide and amygdala functional connectivity profiles

**DOI:** 10.1101/2025.09.15.25335797

**Authors:** Kevin J. Clancy, Caitlin Ravichandran, Sydney A. Jobson, Victor May, Sayamwong E. Hammack, William A. Carlezon, Kerry J. Ressler, Scott L. Rauch, Isabelle M. Rosso

**Author notes:** Correspondence: Kevin J. Clancy, PhD and Isabelle M. Rosso, PhD 115 Mill Street, Mailstop 334, Belmont, MA 02478, and.

## Abstract

Posttraumatic stress disorder (PTSD) is a highly heterogeneous psychiatric disorder, complicating efforts to identify consistent biological markers and develop targeted treatments for individuals exposed to trauma. Recent research has identified a distinct intrusive-hypervigilant (IH) phenotype, which is characterized by heightened intrusive reexperiencing and hypervigilance symptoms along with elevated levels of pituitary adenylate cyclase-activating polypeptide (PACAP), a neuropeptide involved in stress response via amygdala signaling. In an independent sample of 172 symptomatic trauma-exposed adults, we replicated this IH phenotype using latent profile analysis of Clinician-Administered PTSD Scale for DSM-5 symptom severity ratings and expanded its biological characterization using resting-state functional magnetic resonance imaging (rs-fMRI). Consistent with prior work, the identified IH group demonstrated more severe intrusive reexperiencing and hypervigilance symptoms and higher PACAP levels compared to groups with generally High or Low symptom severity. Additionally, the IH phenotype exhibited stronger functional connectivity of the centromedial, but not basolateral, amygdala with regions in the occipital cortex, precuneus, and medial prefrontal cortex - areas primarily within the Default Mode and Visual Networks. Meta-analytic decoding linked these regions to mental imagery, memory processing, fear, and threat perception. These findings support the existence of an IH phenotype of posttraumatic stress that may exhibit a distinct biological profile, characterized by exaggerated interactions between memory, threat, and arousal systems that may be mediated by PACAP and its effects of amygdala connectivity. This phenotype may serve as a promising target for precision psychiatry approaches, including pharmacological and neurotherapeutic interventions that modulate PACAP signaling and amygdala connectivity.

## INTRODUCTION

Posttraumatic stress disorder (PTSD) is a markedly heterogeneous psychiatric disorder, with potentially 636,129 different combinations of 20 symptoms across four distinct symptom clusters that fulfill DSM-5 diagnostic criteria for PTSD [1,2]. Beyond the diagnosis of PTSD, a significant number of trauma-exposed individuals report clinically significant distress and impairment through the endorsement of PTSD symptoms that do not meet full DSM-5 diagnostic criteria (i.e., subthreshold PTSD) [3]. Combined, this vast clinical heterogeneity hinders efforts to identify reliable biological substrates that are needed to advance mechanism-based therapeutics of posttraumatic stress.

The heterogeneity of posttraumatic stress has motivated the identification of “subtypes”, or distinct clinical phenotypes that may have more homogeneous biological mechanisms that can be reliably targeted under a precision psychiatry framework. However, to date, the emergence of reliable subtypes remains elusive. A dissociative subtype of PTSD is the only recognized diagnostic subtype based on DSM-5 criteria and is characterized by the additional endorsement of dissociative symptoms beyond the “classic” 20 PTSD symptoms. Additionally, the ICD-11 recognizes Complex PTSD as a distinct diagnosis that adds disturbances in self-organization to core PTSD symptoms [4]. Beyond these recognized subtypes, additional work has offered evidence for other subtypes based on a predominance of internalizing versus externalizing or fear versus dysphoria symptoms [5–9]. However, other work suggests that the most common subgroups across latent profile and class analyses of PTSD symptoms are “low”, “medium”, and “high” symptoms [10,11].

Importantly, much of this work has utilized samples of participants with shared or similar traumatic experiences. This approach may fail to capture the diverse etiology of PTSD, as different types of traumatic experiences (e.g., single versus recurrent, interpersonal versus non-interpersonal) may result in different symptoms and neurobiological alterations [12,12–15]. Moreover, prior latent class and profile analyses focus on samples meeting full diagnostic criteria for PTSD. This overlooks the significant number of trauma-exposed individuals who experience clinically-significant distress and impairment but do not meet diagnostic thresholds for all four symptom clusters [3].

Addressing these concerns in a diverse, trauma-exposed sample with subthreshold and threshold PTSD, Adams et al. 2025 recently identified a distinct clinical phenotype of posttraumatic stress that was marked by specific elevations in intrusive reexperiencing and hypervigilance symptoms [16]. Moreover, they demonstrated that the intrusive-hypervigilant (IH) clinical profile had impaired fear extinction retention and robust elevations in circulating levels of pituitary adenylate cyclase-activating polypeptide (PACAP). PACAP is a highly-conserved neuropeptide that serves as a critical regulator of stress response, including the activation of physiological arousal and defensive responding through signaling within the amygdala complex, particularly the central amygdala [17–24]. It has been linked to the pathophysiology of PTSD through alterations in amygdala activity and subsequent effects on arousal, threat processing, and fear memory [25,26]. Recent evidence further links PACAP to exaggerated functional connectivity of the amygdala with regions of the default mode network (DMN), a network frequently implicated in PTSD through its role in autobiographical memory, arousal, and internally (versus externally) oriented attention [27]. Additional evidence suggests that PACAP’s effects are stronger in female than male patients, indicating that it may reveal mechanistic insights into sex differences in PTSD risk and prevalence [16,25]. Correspondingly, Adams et al. demonstrated that the IH subtype with elevated PACAP was predominantly female, suggesting a sex-specific biological phenotype of posttraumatic stress. Overall, their findings identify a potentially more homogenous clinical-biological subgroup that pairs elevated intrusive reexperiencing and hypervigilance symptoms with biological and psychophysiological markers of fear memory and arousal. This may represent an important step towards a biologically-grounded clinical subtype to target through mechanism-based interventions in a precision psychiatry approach.

The current study sought to replicate findings of an IH clinical phenotype with elevations in PACAP and build upon their biological profile by providing novel evidence for distinct patterns of amygdala functional connectivity. Using resting-state functional magnetic resonance imaging (rs-fMRI) in an independent sample of symptomatic trauma-exposed adults, we tested the hypothesis that an IH phenotype would be associated with exaggerated functional connectivity of the amygdala with networks implicated in trauma memory and arousal. Consistent with prior work, we hypothesized these effects would be specific to the right central amygdala, where PACAP-resting state associations are strongest [27].

## METHODS

### Participants

One-hundred and seventy-two trauma-exposed adults were recruited and enrolled via advertisements in the local community. Study procedures were approved by the Mass General Brigham Human Research Committee and all participants provided written informed consent. Participants completed a fasting blood draw, clinical interview, self-report questionnaires, and a 13-minute eyes-open resting-state fMRI scan. Participants were included if they met DSM-5 diagnostic criteria for at least two PTSD symptom clusters based on the Clinician Administered PTSD Scale for DSM-5 (CAPS-5), constituting subthreshold (n = 57; 33%) and threshold (n = 115; 67%) PTSD [3]. Given a priori interest in biological sex differences motivated by prior work [28], participants were required to be the same sex as assigned at birth, female participants were to be premenopausal, and participants with a history of receiving hormonal replacement therapy or undergoing surgery to change biological sex were excluded. Other exclusion criteria are detailed in Clancy et al. 2023 [27].

Of the 172 enrolled participants with complete clinical interview data, 158 (106 female) had detectable serum PACAP levels (excluded: undetectable values = 13, levels below reliable detection threshold = 1). A total of 134 (85 female) had usable fMRI data (excluded: excessive motion = 14, incomplete scanning = 7, poor structural-functional alignment = 4, significant artifact = 6, no scanning performed = 7). Demographic and clinical characteristics of the sample are provided in **Table 1**.

**Table 1.**
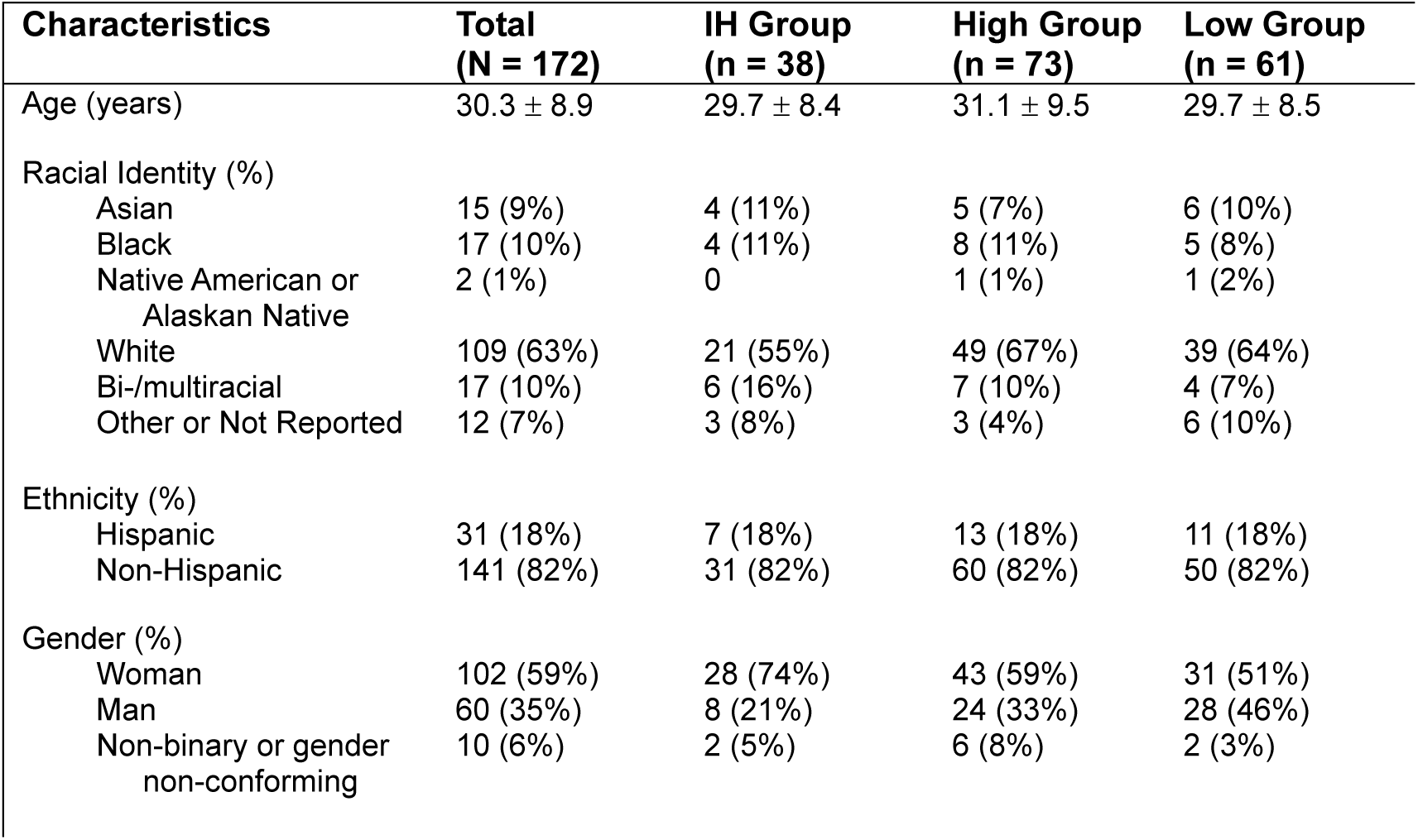
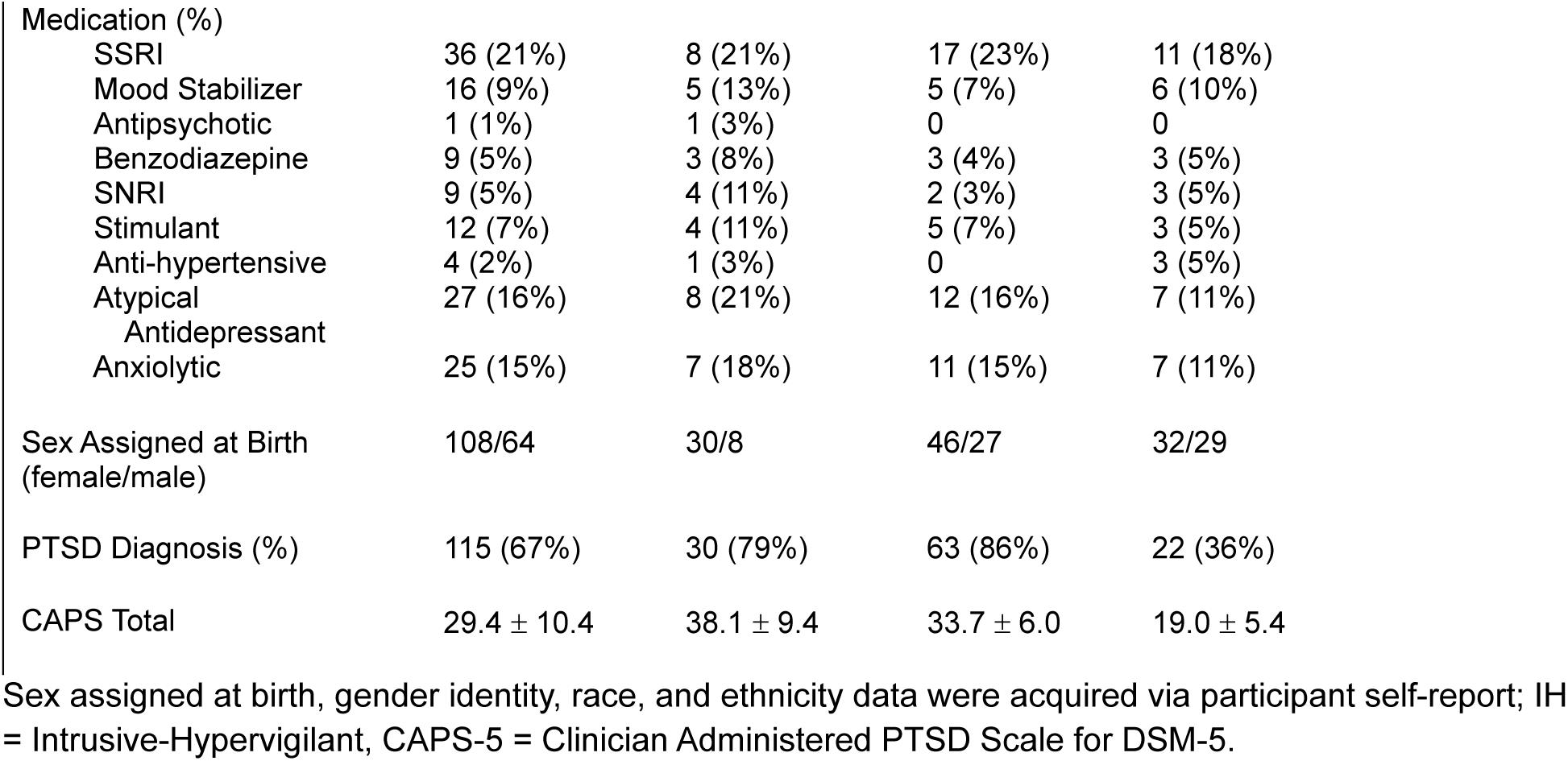
Demographic and clinical characteristics of the sample. Means ± standard deviations or N (%).

### Clinician Administered PTSD Scale for DSM-5 (CAPS-5)

The CAPS-5 [29], the gold-standard diagnostic interview for PTSD, was administered by doctoral-level clinicians. This interview consists of 30 items designed to assess the onset, duration, and impact of PTSD symptoms, yielding a determination of PTSD diagnosis and symptom severity. Specifically, each of the 20 DSM-5 PTSD symptoms are given an overall severity rating, which reflects the intensity and frequency of the symptom within the past month. Severity ratings for each symptom are made using a 0-4 Likert Scale (0 = Absent, 4 = Extreme/incapacitating).

### PACAP

Details about PACAP collection and analysis are provided in Clancy et al. 2023 [27]. Briefly, PACAP38-specific immunoassays (Cat. No. HUFI02692, AssayGenie, Dublin, Ireland) were performed at the University of Vermont - Larner College of Medicine using human plasma samples that were collected at the beginning of each study visit, between approximately 8:00- 10:00AM. Fourteen samples were excluded due to undetectable (n = 13) or unreliably low (n = 1) PACAP levels, resulting in a final sample of n = 158 for PACAP analyses. Consistent with Adams et al. 2025, PACAP values were square-root-transformed to reduce data compression and correct a left-skewed distribution. PACAP levels were analyzed in two batches. There was a significant effect of Batch on PACAP levels (Batch 1 mean = 29.53, Batch 2 mean = 20.98; *t* = 3.08, *p* = 0.003), with unequal variances across the two batches (Leven’s Test F = 6.22, *p* = 0.014). These Batch differences were not explained by clinical or demographic variables; therefore, Batch (binary 1 or 2) was included as a covariate in all PACAP analyses.

### Latent Symptom Profile Analysis

Gaussian mixture modeling was performed using the Mclust package in R [30] to identify latent symptom profiles using symptom severity ratings from the 20-item CAPS-5 interview. Specifically, we aimed to replicate the IH symptom profile identified in Adams et al. 2025 through a complimentary yet distinct approach to demonstrate the external reliability of this symptom profile in an independent sample. Four models with different covariance structures were examined, each testing a range of 1 to 4 profiles (as in, Adams et al. 2025), resulting in a total of 16 models examined. In two of the models, clustered profiles were assumed to have equal volume and shape, with one model having an equal diagonal covariance matrix (EEI model) and the other allowing for full covariance matrices (EEE model) across profiles. The other two models permitted variable volumes and shapes across profiles, with one utilizing an equal diagonal covariance matrix (VVI model) and the other allowing full covariance matrices (VVV model) across profiles. Optimal model fit was determined using Bayesian Information Criterion (BIC), comparing BIC values for all 16 different model fits.

### MRI data acquisition and preprocessing

Imaging was conducted at the McLean Hospital Imaging Center on a 3T Siemens Prisma scanner with a 64-channel head coil using Human Connectome Project (HCP) sequences for anatomical and functional images. MRI data were preprocessed using fMRIPrep version 20.2.7 [31] with non-aggressive AROMA denoising of motion artifacts [32]. Participants were excluded if their mean framewise displacement (FD) exceeded 0.5mm or greater than 20% of volumes exceeded FD = 0.5 mm (n = 14). Subsequent denoising of residual physiological confounds from white matter and cerebrospinal fluid using the CompCor method, scrubbing of motion outliers (FD > 0.5mm), and high pass (0.1 Hz) filtering were performed using CONN [33]. In addition to the 14 excluded from analysis for excessive motion, 7 had incomplete scanning, 4 had inadequate alignment of structural and functional scans, 6 had significant imaging artifact, and 7 had no scanning performed, resulting in a final sample of n = 136 for fMRI analyses. Protocol details and additional denoising steps are reported in Clancy et al. 2023 [27].

### Resting-state functional connectivity analyses

Seed-based resting state connectivity analyses were conducted in CONN using cleaned timeseries of BOLD signal extracted from left and right (l/r) centromedial (CMA) and basolateral (BLA) amygdala regions of interest (ROIs) generated from the JuBrain Atlas [34,35]. Note that the JuBrain Atlas combines both central and medial AMYG nuclei into a single CMA ROI, as segmentation of these adjacent subnuclei has been unreliable in human 3T fMRI (Figure S1). Whole-brain seed-based correlations were performed for l/rCMA and l/rBLA seeds to compute whole-brain functional connectivity (FC) maps. Seed-based FC values were Fisher’s z-transformed prior to further statistical analyses.

### Analytic Plan

Given our specific *a priori* interest in an IH symptom profile, an identified group matching this profile was compared to other symptom groups to determine the clinical and biological correlates of this IH phenotype. First, the identified IH group was compared to all other symptom profiles combined into a single non-IH group, as prior findings showed no PACAP differences between non-IH clinical phenotypes (Adams et al. 2025). This approach was contingent on no statistical differences existing between the non-IH clinical phenotypes across the biological markers of interest (i.e., PACAP levels and amygdala FC) in our sample. Comparisons of the IH group versus the single non-IH group were followed by separate comparisons between the IH group and other groups individually.

Group comparisons were examined using Chi-squared tests of proportions for sex assigned at birth (male vs. female) and PTSD diagnosis (yes vs. no) using the prop.test function in R [36], testing for higher female representation and PTSD diagnosis rates in the IH group.

Model-based estimated marginal means in PACAP levels by phenotypic group, and test statistics and p-values for associated significance tests, were calculated from results of linear regression models controlling for Batch using the emmeans package for R [37]. Robust standard errors were used for all analyses examining PACAP with Batch as a covariate due to unequal variances the batches. T-test contrasts examined Group differences in whole-brain seed-based amygdala FC maps. Whole-brain FC results were thresholded at p < 0.005 (uncorrected height threshold), p < 0.0125 FDR-corrected cluster threshold, to correct for multiple comparisons across the 4 amygdala ROIs (0.05/4 ROIs = 0.0125). The SPM Anatomy Toolbox was used to assign estimated anatomical labels for significant clusters [38]. Meta-analytic decoding of identified clusters was performed using the NiMARE v0.0.11 package which ascribes functional labels to images using the NeuroSynth meta-analytic database [39–42].

Finally, we tested associations between clinical symptoms and biological markers. Individual linear regressions controlling for Batch examined associations between PACAP levels and symptoms that differentiated the IH group. Pearson correlations examined associations between symptom severity and amygdala functional connectivity.

One-tailed tests were used for replication analyses (e.g., elevated PACAP in IH), and two-tailed tests for novel analyses (e.g., functional connectivity differences between groups, symptom associations with identified biological markers). One-sided p-values are explicitly labeled in the results; all other p-values are two-sided.

## RESULTS

### Latent Profile Analysis

Consistent with Adams et al. 2025, a three-profile EEI model of CAPS-5 symptoms provided the best fit based on the BIC. The resulting profiles included: a High group (n = 73) characterized by elevated severity across most PTSD symptoms; a Low group (n = 61) with generally low symptom severity; and an Intrusive-Hypervigilant (IH) group (n = 38) marked by differential elevations in intrusion and hypervigilance symptoms. Compared to the High and Low groups, the IH Group demonstrated significantly greater severity of trauma-related intrusive memories (B1; IH > High/Low: t = 3.04/6.38, p = 0.008/< 0.0001), flashbacks (B3; IH > High/Low: t = 34.17/33.55, p’s < 0.0001), and hypervigilance (E3; IH > High/Low: t = 2.87/4.27, p = 0.013/0.0001; **Figure 1**).

**Figure 1.**
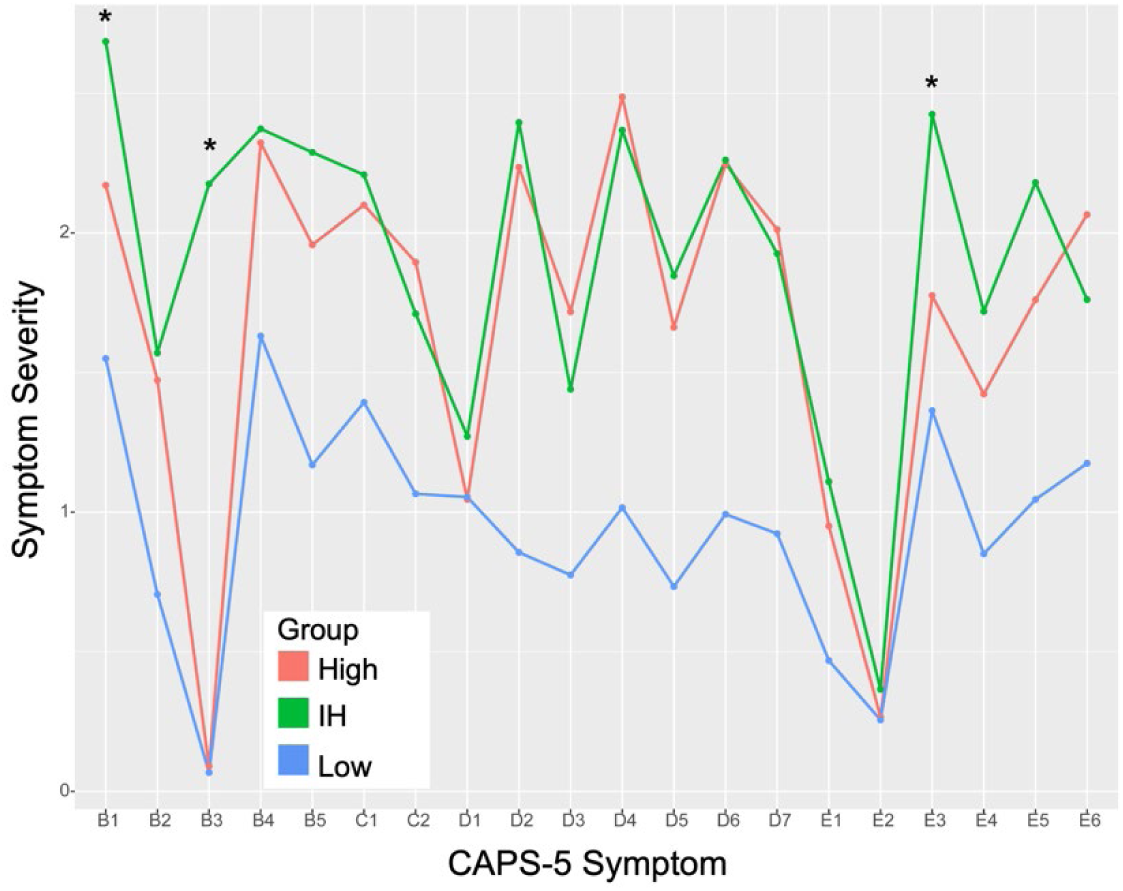
CAPS-5 symptom profiles of the groups identified from the latent profile analysis. * indicates where the Intrusive-Hypervigilant (IH) group differed from both the High and Low groups. B = Intrusive Reexperiencing symptoms (Cluster B); C = Avoidance symptoms (Cluster C); D = Negative Alterations in Cognitions and Mood symptoms (Cluster D); E = Hyperarousal symptoms (Cluster E).

Also in line with prior findings, the IH group had a greater proportion of females (79%) compared to the High (63%; IH > High χ^2^ = 2.94, *p* = 0.043) and Low groups (52%; IH > Low χ^2^ = 7.02, *p* = 0.004). The High and Low groups did not differ in sex composition (High ≠ Low χ^2^ = 2.94, *p* = 0.217). We also observed differences in the proportion of threshold (vs. subthreshold) PTSD diagnoses, with the IH group having a greater proportion (79%) compared to the Low group (36%; IH > Low χ^2^ = 17.27, *p* < 0.001) but not the High group (86%; IH > High χ^2^ = 0.99, *p* = 0.841). The High group also had significantly more full PTSD cases than the Low group (High > Low χ^2^ = 36.16, *p* < 0.001).

### PACAP38

There were no significant differences in PACAP levels between the High (M = 24.02, SE = 1.63) and Low groups (M = 22.86, SE = 1.77; *t*(120) = 0.52, *p* = 0.607), justifying their combination into a single non-IH comparison group. The IH group (M = 29.08, SE = 2.53) had significantly higher PACAP levels compared to the combined non-IH group (*t*(155) = 1.97, *p* = 0.025 one-tailed, *d* = 0.39). Follow-up contrasts confirmed elevated PACAP levels in the IH group compared to both the High (*t*(154) = 1.65, *p* = 0.050 one-tailed, *d* = 0.35) and Low groups (*t*(154) = 1.98, *p* = 0.025 one-tailed, *d* = 0.44; **Figure 2**).

**Figure 2.**
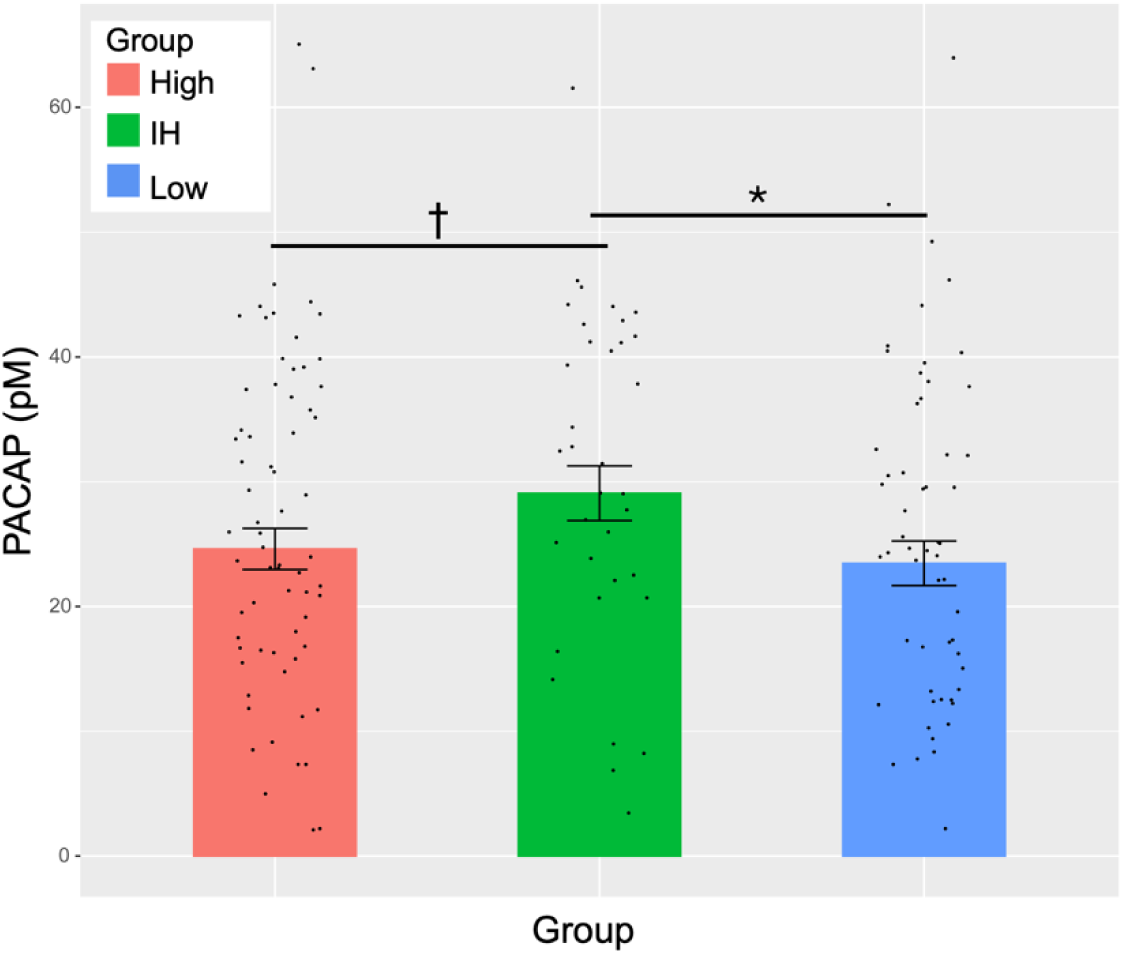
Differences in circulating PACAP levels across participants with generally high (High), generally low (Low), and selectively elevated Intrusive-Hypervigilant (IH) PTSD symptom profiles. Bars represent estimated marginal means from linear regression models controlling for batch effects; error bars reflect standard errors from the mean. Individual points reflect raw (untransformed) PACAP values. Data. PACAP values were square-root transformed for statistical analysis. † *p* = 0.05 one-tailed; * *p* < 0.05 one-tailed.

### Functional Connectivity

Whole-brain rCMA-seed connectivity maps confirmed no differences between the High and Low groups, again supporting their combination into a single group. Relative to this combined High/Low group, the IH group exhibited stronger rCMA connectivity with three midline clusters (**Figure 3A**): a cluster spanning the parietooccipital sulcus and intracalcarine cortex (k = 302 voxels, cluster FDR *q* = 0.001, peak MNI = [-4, -72, 16], *T* = 4.15), a precuneus cortex cluster (k = 287, cluster FDR *q* = 0.001, peak = [-4, -56, 68], *T* = 4.73), and a medial prefrontal cortex (mPFC) cluster comprising the paracingulate gyrus and anterior cingulate cortex (ACC; k = 210, cluster FDR *q* = 0.005, peak = [-2, 46, 14], *T* = 4.63). Mapping these clusters onto Yeo’s 7-network atlas (**Figure 3B**) situated them predominantly within the Default Mode Network (k = 212) and Visual Network (k = 208), and to a lesser extent the Dorsal Attention (k = 99), Ventral Attention (k = 81), Somatomotor (k = 76), and Frontoparietal (k = 46) networks. Follow-up group contrasts revealed the IH group demonstrated greater rCMA FC across all three clusters compared to both the High and Low groups (**Figure 3C; Supplemental Table 1**). No significant group differences emerged for seed connectivity from the rBLA, lCMA, or lBLA.

**Figure 3.**
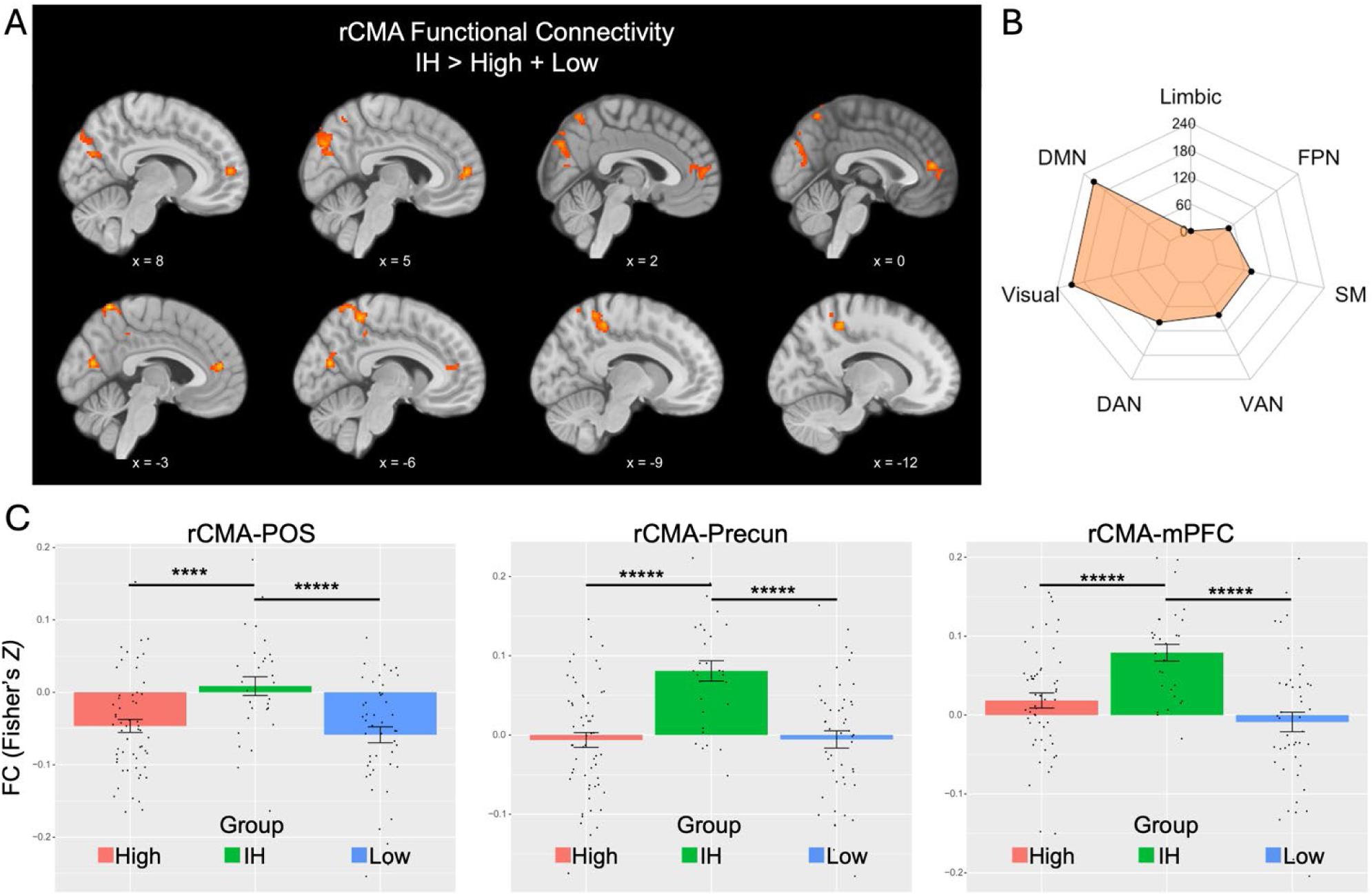
Group differences in right centromedial amygdala (rCMA) resting-state functional connectivity (FC). A) Whole-brain contrasts (IH > non-IH) revealed three significant clusters showing greater rCMA FC in the Intrusive-Hypervigilant (IH) group within the parietooccipital sulcus (POS), precuneus (Precun), and medial prefrontal cortex (mPFC). B) Radar plot depicting the number of voxels within the significant clusters that overlap with the 7 networks of the Yeo atlas, demonstrating the clusters were predominantly within the default mode network (DMN) and Visual network. C) Post hoc individual group contrasts revealed that the IH group showed significantly greater rCMA FC than both the High and Low symptom groups across all three clusters. FPN = frontoparietal network; SM = somatomotor network; VAN = ventral attention network; DAN = dorsal attention network; **** < 0.001, ***** < 0.0005, ****** < 0.0001.

### Associations with clinical symptoms

Across all participants, PACAP levels were modestly associated with CAPS-5 item severity scores for flashbacks (β = 0.17, SE = 0.08, *p* = 0.025; **Figure 4A**) but not trauma-related intrusive memories (β = 0.08, SE = 0.08, *p* = 0.336) or hypervigilance (β = 0.02, SE = 0.08, *p* = 0.787). Flashback severity was additionally associated with rCMA-POS (*r* = .35, *p* < 0.0001; **Figure 4B**), rCMA-Precun (*r* = 0.43, *p* < 0.001; **Figure 4C**), and rCMA-mPFC FC (*r* = 0.37, *p* < 0.0001; **Figure 4D**). Additional weak and trending associations were seen between rCMA-mPFC FC and hypervigilance (*r* = 0.17, *p* = 0.048), as well as intrusive memories (*r* = 0.15, *p* = 0.079; **Figure 4D**).

**Figure 4.**
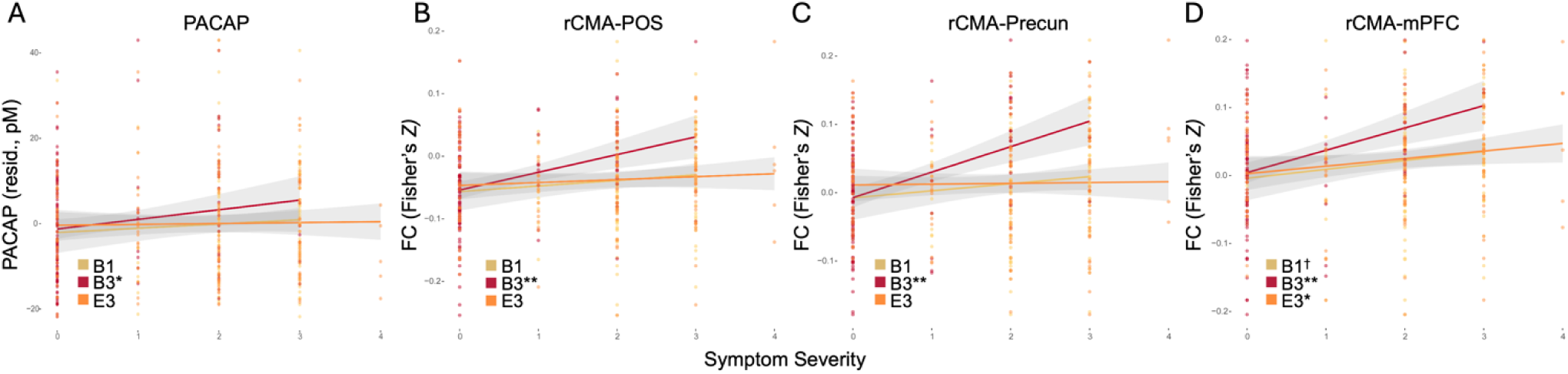
Associations between posttraumatic stress disorder (PTSD) symptom severity and biomarkers of interest. **A.** Pituitary adenylate cyclase-activating (PACAP) levels (square-root-transformed) were significantly associated with flashback severity (CAPS-5, item B3) but not with trauma-related intrusive memories (B1) or hypervigilance (E3). **B–D.** Right centromedial amygdala (rCMA) functional connectivity with the parieto-occipital sulcus (POS), precuneus (Precun), and medial prefrontal cortex (mPFC) was positively associated with flashback severity. Weaker, trending associations were also observed between rCMA–mPFC connectivity and both intrusive memories and hypervigilance. CAPS-5: Clinician-Administered PTSD Scale for DSM-5.

## DISCUSSION

In an independent sample of trauma-exposed adults with symptoms spanning subthreshold to threshold DSM-5 PTSD, we replicated the findings of Adams et al. 2025, identifying an IH clinical phenotype characterized by more severe intrusive memories, flashbacks, and hypervigilance, as well as elevated levels of circulating PACAP. Extending prior work, we provide novel evidence that this IH phenotype is associated with stronger resting state FC of the rCMA with midline cortical regions, particularly within the default mode and visual networks. Importantly, both PACAP levels and rCMA FC were significantly associated with more severe flashback symptoms, supporting their potential as biomarkers for this clinical phenotype.

Our identification of an IH clinical phenotype aligns with existing neurocognitive models of PTSD symptoms that highlight the intersection of arousal, memory, and attention. The “warning signal” hypothesis of intrusive trauma reexperiencing symptoms posits that neutral sensory cues can elicit exaggerated arousal and trigger intrusive memories or flashbacks through acquired temporal associations with the traumatic event [43]. This conditioned threat to neutral sensory cues one encounters in everyday life may lead to exaggerated externally-oriented attention to detect and respond to such threat cues – the hallmark presentation of hypervigilance.

Translational evidence suggests that “hypervigilance” and its proposed neurochemical underpinnings increase sensory sensitivity, reduce sensory gating, and facilitate rapid sensory processing, potentially through alterations in both top-down and bottom-up attention [44–53]. Therefore, hypervigilance may contribute to the detection and rapid processing of sensory cues that trigger intrusive trauma memories and flashbacks through the activation of sensory-bound representations of the traumatic event. Moreover, physiological arousal may further exacerbate this cycle, as inductions of physiological arousal can elicit flashback-like memories [54–56].

Taken together, these findings suggest that the IH phenotype represents a clinical presentation of posttraumatic stress driven by interactions between arousal, memory, and attention systems that amplify specific reexperiencing symptoms.

In keeping with this notion, the observed elevations in circulating PACAP levels within the IH phenotype align with the demonstrated role of PACAP in stress physiology and fear memory.

PACAP is a critical regulator of arousal through its signaling in autonomic stress pathways [18,19,57]. PACAP and its predominant type 1 receptor (PAC1R) are densely expressed within the central extended amygdala [17–20], encompassing both the central amygdala (CeA) and bed nucleus of the stria terminalis (BNST), which activate fear and panic reflexes in response to threat through downstream projections to physiological arousal systems [58]. PACAP is also linked to the acquisition, consolidation, and extinction of contextual or cue-based fear memory through alterations in synaptic plasticity within fear memory circuits [59–63]. Notably, PACAP signaling facilitates trace fear memory, a form of fear learning that is dependent on sustained attention and episodic memory systems [64] and is highly aligned with the “warning signal” model of intrusive reexperiencing symptoms. To this end, PACAP may facilitate attentional states of hypervigilance for threat as a consequence of sustained conditioned fear. Moreover, PACAP may enhance consolidation and impair extinction of contextual fear memories that are linked to episodic memory systems, akin to trauma-related intrusive memories and flashbacks. Aligning with the neurocognitive models of PTSD, this cycle may be maintained and exacerbated by the elevated states of physiological arousal driven by PACAP signaling. Therefore, PACAP is optimally positioned at the intersection of arousal, memory, and attention systems to give rise to the IH phenotype.

Strengthened functional connectivity between the amygdala and the identified posterior and prefrontal regions further supports a role of arousal-memory-attention interactions. Stronger functional coupling between the amygdala and precuneus is associated with greater threat reactivity [65], hyperarousal [27], and sustained attention to aversive stimuli [66]. Additionally, both the amygdala and our identified precuneus cluster are linked to the retrieval of emotional autobiographical memory [67–70]. Our identified mPFC cluster overlaps with the pregenual ACC (Brodmann’s Area 32), whose rodent analogue, the prelimbic cortex, is consistently implicated in the expression of conditioned fear through connectivity with the amygdala [71–73]. Specifically, the retrieval of long-term conditioned fear memory may be supported by strengthened connections between the prelimbic cortex and central amygdala through the paraventricular thalamus [74,75]. In humans, the pregenual ACC is engaged during the retrieval of remote contextual fear memories that involve the amygdala and hippocampus [76–79]. Similarly, the POS and adjacent visual cortex have been tied to the retrieval of vivid sensory details of episodic memories [80–83] through their role in visual imagery and visuospatial processing [84,85]. More broadly, these posterior cortices are involved in emotional arousal [86], vigilance [87], and directed attention [88]. In sum, while these regions support numerous functions, they converge on a pattern of processes that are tied to the detection, processing, and responding to threat and the consolidation and retrieval of emotional episodic or fear memories. Our meta-analytic decoding of this collective pattern of posterior and prefrontal clusters provides evidence for this convergence, as it revealed a spatial overlap with studies ascribing functions such as mental imagery, memory retrieval and encoding, negative emotions, fear and threat, and attention (**Supplemental Table 2**).

Collectively, these identified regions fall predominantly within the DMN and Visual Network (**Figure 3B**). Both networks are consistently implicated in posttraumatic stress given their respective functions in episodic memory, attention, and arousal [89–95]. Typically, these networks demonstrate robust anticorrelation and are positioned at opposite ends of the hierarchical organization of cortical networks, reflecting a maximal segregation in their patterns of activity and connectivity [39]. However, there is accruing evidence for a breakdown in the functional segregation of these intrinsic networks in posttraumatic stress [96,97], particularly in relation to symptoms of hypervigilance [91] and trauma reexperiencing [98]. Additionally, an imbalance in their anticorrelation, or co-deactivation, has been linked to the sensory-perceptual features of trauma memories, including their emotional intensity and sense of reliving which are key features of flashbacks [81]. The current findings of strengthened connectivity between the amygdala and central brain regions of both the DMN and Visual Network may similarly reflect increased functional integration of these normally divergent networks. This suggests that the IH clinical phenotype is marked by abnormal coupling between these intrinsic resting state networks and the amygdala, reflecting a potential biomarker for this subtype. Future work incorporating dynamic functional connectivity of these networks are needed to elucidate the network-wide versus region-specific alterations and which patterns maximally differentiate the IH phenotype. Moreover, task-based neuroimaging may be able to determine whether these changes in organization are in fact tied to behavioral measures of arousal, memory, and attention.

Interestingly, our functional connectivity findings were specific to the rCMA, as no effects emerged with the lCMA or r/lBLA. The BLA generally demonstrates more robust functional connectivity with cortical structures compared to the CMA, which preferentially connects with subcortical structures like the midbrain and striatum [99–102]. Our findings replicate this pattern, as the non-IH groups demonstrated near-zero average connectivity between the CMA and cortical regions (Figure 3C). Prior work has demonstrated increased BLA-cortical connectivity in PTSD compared to trauma-exposed controls, but no differences in CMA connectivity due potentially to the minimal intrinsic CMA-cortical connectivity [99]. However, we previously demonstrated that elevated levels of circulating PACAP are associated with strengthened cortical connectivity of the rCMA, and not the BLA, with midline regions of the DMN, including the precuneus [27]. Our present findings qualify this unusual strengthening of CMA-cortical connectivity by demonstrating a specificity of this effect to the IH clinical phenotype, which was the only clinical phenotype with elevations in PACAP levels. Therefore, this abnormal rCMA connectivity pattern may reflect a PACAP-mediated shift in the functional organization of the amygdala and its cortical interactions that is specific to the IH phenotype. These converging findings position the combination of high PACAP and rCMA functional connectivity as a candidate biomarker for the IH clinical phenotype, reflecting a distinct neurobiological profile not observed in other posttraumatic stress presentations.

Critically, the High and Low symptom groups did not differ in PACAP levels or rCMA functional connectivity despite notable differences in nearly every symptom, including intrusive memories and hypervigilance (Figure 1). This specificity reinforces the hypothesis that the IH phenotype may reflect a biologically distinct subtype of posttraumatic stress, characterized clinically by differentially elevated intrusive memories, flashbacks, and hypervigilance. Among these, flashbacks emerged as the only individual symptom significantly associated with PACAP and rCMA connectivity. This may be attributable to the greater physiological arousal and cognitive alterations often associated with flashbacks compared to other trauma-related intrusive memories, reflecting an exaggerated interaction among memory, attention, and arousal systems [103]. Neuroimaging studies examining the reliving or re-enacting of traumatic events further emphasize a coupling between the DMN and sensory cortex in flashbacks, reflecting a reorganization of intrinsic connectivity patterns that is marked by the integration of sensory processing and autobiographical memory retrieval [96]. Notably, flashbacks were also the most robust differentiator between the IH and non-IH groups. These findings suggest that flashbacks may serve as a core clinical marker of the distinct biological profile underlying the IH phenotype. External replications with independent trauma-exposed samples are needed to confirm this specificity and validate these biomarkers of an IH phenotype, including the potential centrality of flashbacks symptoms.

The present findings are not without limitations. The cross-sectional design precludes inference of directionality in associations between PACAP, rCMA connectivity, and clinical phenotypes. Longitudinal and experimental studies are needed to gain insights into the temporal relationship between these biological and clinical markers to better determine if elevated PACAP and rCMA connectivity are biological mechanisms of the IH phenotype, or mere physiological sequalae of symptoms. Relatedly, all imaging data were collected at rest; as such, we cannot directly support claims linking PACAP and rCMA-cortical connectivity to the active processes of memory, attention, and arousal that are implicated in intrusive reexperiencing and hypervigilance symptoms. Future studies incorporating cognitive testing or behavioral measures of these constructs in conjunction with task-based neuroimaging are needed to further clarify the mechanistic processes giving rise to the IH phenotype. Finally, our functional connectivity analyses were constrained to amygdala subregions, specifically the BLA and CMA, given prior work demonstrating robust PACAP signaling within these regions. However, PACAP is expressed in numerous other subcortical and midbrain regions that are tied to behavioral arousal, attention, and memory [18]. Many of these structures require greater anatomical precision to delineate them from adjacent structures and sources of physiological noise. Future studies utilizing ultra-high-field MRI (i.e., 7T) or specialized sequences (e.g., neuromelanin-sensitive imaging) would afford deeper insights into other subcortical structures that may be mechanistically linked to the IH phenotype.

In conclusion, this study replicates the identification of an intrusive-hypervigilant clinical phenotype of posttraumatic stress that is characterized by elevated circulating PACAP levels and provides novel evidence for altered patterns of functional connectivity of the rCMA. This IH phenotype may reflect a distinct clinical presentation that is characterized by exaggerated interactions between memory and arousal processes, thus driving specific symptoms that are situated at this intersection, like flashbacks. Our findings further add to the accumulating evidence linking both PACAP and altered organization of subcortical-cortical networks to memory, arousal, and PTSD. More specifically, the IH phenotype exhibits these biomarkers previously implicated in the PTSD literature, which have been stymied by inconsistent replication across different, diverse samples. Therefore, the IH phenotype may be a candidate clinical presentation for a precision psychiatry framework that targets these biomarkers through pharmacotherapy or neurotherapeutics.

## Data Availability

All data produced in the present study are available upon reasonable request to the authors

## Acknowledgments

The authors would like to thank all participants for dedicating their time and energy to completing the study. We also thank the MRI technologists of the McLean Imaging Center.

## Author contributions

KJC performed data processing, statistical analyses, and wrote the initial draft of the manuscript. CR provided consultation on all statistical analyses and provided critical feedback in the analytic plan and reporting of the statistical methods and results. VM and SEH performed analysis of PACAP data. SAJ contributed to data collection, management, and processing. WAC, KJR, SLR, and IMR obtained funding and contributed to study design and conceptualization. SLR and IMR conceived the study. All authors provided critical input to the manuscript, contributed to revised drafts, and approved the final manuscript.

## SUPPLEMENT

**Figure S1.**
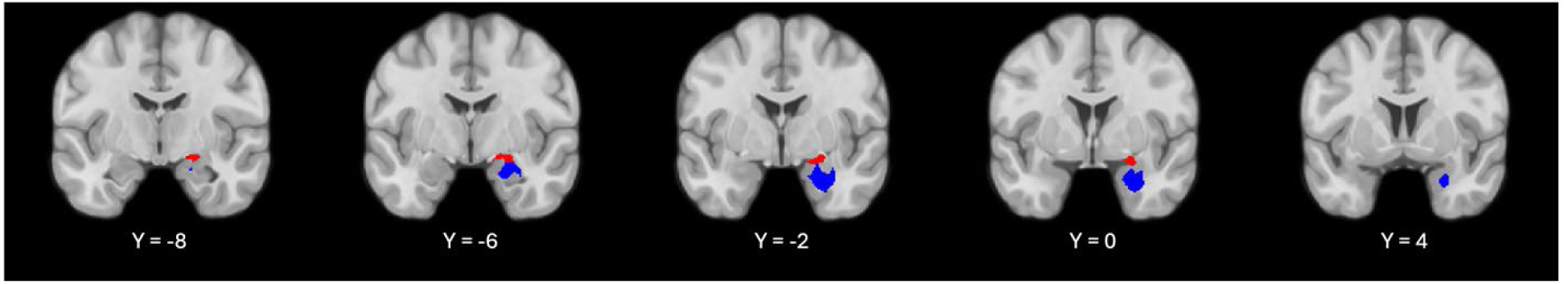
Amygdala subregions of interest. Anatomical masks of the centromedial (red) and basolateral (blue) amygdala subregions used for functional connectivity analyses. Masks were generated from the JuBrain Atlas.

**Supplemental Table 1.** Individual CAPS-5 symptom group contrasts of resting-state rCMA FC.

|  | IH vs. High | IH vs. Low | High vs. Low |
| --- | --- | --- | --- |
| rCMA-POS | $t = 3.43, p = 0.0008$ | $t = 4.05, p = 0.0001$ | $t = 0.87, p = 0.385$ |
| rCMA-Precuneus | $t = 5.38, p < 0.0001$ | $t = 5.15, p < 0.0001$ | $t = 0.05, p = 0.960$ |
| rCMA-mPFC | $t = 3.60, p = 0.0004$ | $t = 5.04, p < 0.0001$ | $t = 1.85, p = 0.067$ |

rCMA = right centromedial amygdala, POS = parietooccipital sulcus, mPFC = medial prefrontal cortex.

**Supplemental Table 2.**
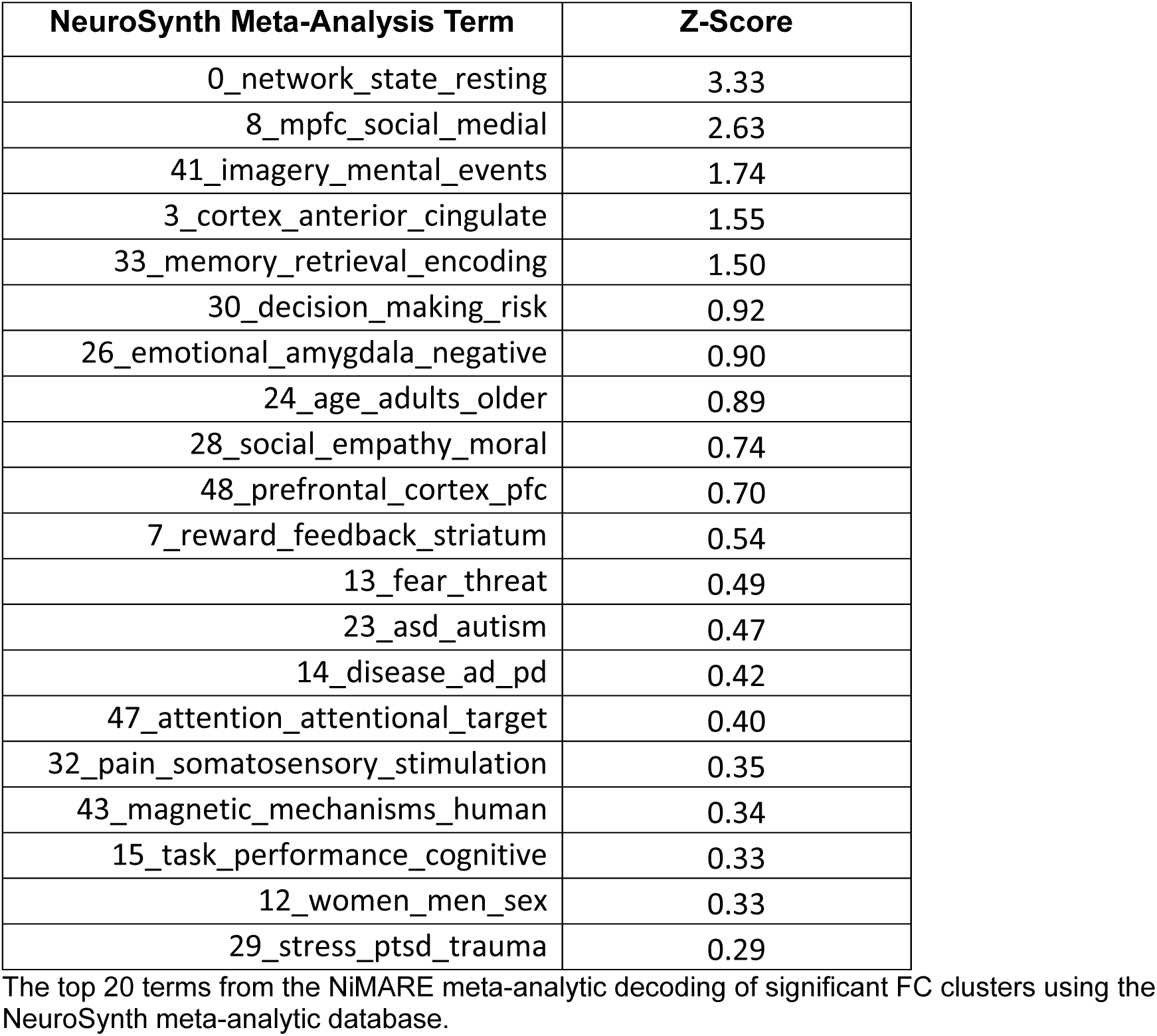
Meta-analytic decoding of significant FC clusters.

## REFERENCES

1. Bryant RA, Galatzer-Levy I, Hadzi-Pavlovic D. The Heterogeneity of Posttraumatic Stress Disorder in DSM-5. JAMA Psychiatry. 2023;80:189–191.

2. Galatzer-Levy IR, Bryant RA. 636,120 Ways to Have Posttraumatic Stress Disorder. Perspect Psychol Sci. 2013;8:651–662.

3. McLaughlin KA, Koenen KC, Friedman MJ, Ruscio AM, Karam EG, Shahly V, et al. Sub- threshold Post Traumatic Stress Disorder in the WHO World Mental Health Surveys. Biol Psychiatry. 2015;77:375–384.

4. Cloitre M, Garvert DW, Brewin CR, Bryant RA, Maercker A. Evidence for proposed ICD-11 PTSD and complex PTSD: a latent profile analysis. European Journal of Psychotraumatology. 2013;4:20706.

5. Campbell SarahB, Trachik B, Goldberg S, Simpson TracyL. Identifying PTSD symptom typologies: A latent class analysis. Psychiatry Research. 2020;285:112779.

6. Forbes D, Elhai JD, Miller MW, Creamer M. Internalizing and externalizing classes in posttraumatic stress disorder: A latent class analysis. Journal of Traumatic Stress. 2010;23:340–349.

7. Miller MW, Kaloupek DG, Dillon AL, Keane TM. Externalizing and Internalizing Subtypes of Combat-Related PTSD: A Replication and Extension Using the PSY-5 Scales. Journal of Abnormal Psychology. 2004;113:636–645.

8. Miller MW, Resick PA. Internalizing and Externalizing Subtypes in Female Sexual Assault Survivors: Implications for the Understanding of Complex PTSD. Behavior Therapy. 2007;38:58–71.

9. Zoellner LA, Pruitt LD, Farach FJ, Jun JJ. Understanding Heterogeneity in Ptsd: Fear, Dysphoria, and Distress. Depression and Anxiety. 2014;31:97–106.

10. Sanghvi DE, Rackoff GN, Newman MG. Latent class analysis of post-traumatic stress disorder symptoms following exposure to Hurricane Ike. Soc Sci Med. 2023;327:115942.

11. Whiteman SE, Lee DJ, Kramer LB, Petri JM, Weathers FW. Subgroup differences in PTSD symptom presentations: A latent class analysis. European Journal of Trauma & Dissociation. 2024;8:100430.

12. Kelley LP, Weathers FW, McDevitt-Murphy ME, Eakin DE, Flood AM. A comparison of PTSD symptom patterns in three types of civilian trauma. Journal of Traumatic Stress. 2009;22:227–235.

13. Lewis MW, Jones RT, Davis MT. Exploring the impact of trauma type and extent of exposure on posttraumatic alterations in 5-HT1A expression. Transl Psychiatry. 2020;10:237.

14. Lewis SJ, Arseneault L, Caspi A, Fisher HL, Matthews T, Moffitt TE, et al. The epidemiology of trauma and post-traumatic stress disorder in a representative cohort of young people in England and Wales. The Lancet Psychiatry. 2019;6:247–256.

15. Smith HL, Summers BJ, Dillon KH, Cougle JR. Is worst-event trauma type related to PTSD symptom presentation and associated features? Journal of Anxiety Disorders. 2016;38:55–61.

16. Adams SW, Neylan TC, May V, Hammack SE, Ressler K, Inslicht SS. PACAP associated with precise PTSD and fear extinction response in women. Psychoneuroendocrinology. 2025;173:107375.

17. Boucher MN, May V, Braas KM, Hammack SE. PACAP orchestration of stress-related responses in neural circuits. Peptides. 2021;142:170554.

18. Hammack SE, May V. Pituitary adenylate cyclase activating polypeptide in stress-related disorders: data convergence from animal and human studies. Biol Psychiatry. 2015;78:167–177.

19. Hashimoto H, Shintani N, Tanida M, Hayata A, Hashimoto R, Baba A. PACAP is implicated in the stress axes. Curr Pharm Des. 2011;17:985–989.

20. Missig G, Roman CW, Vizzard MA, Braas KM, Hammack SE, May V. Parabrachial nucleus (PBn) pituitary adenylate cyclase activating polypeptide (PACAP) signaling in the amygdala: implication for the sensory and behavioral effects of pain. Neuropharmacology. 2014;86:38–48.

21. Patel R, Spreng RN, Shin LM, Girard TA. Neurocircuitry models of posttraumatic stress disorder and beyond: a meta-analysis of functional neuroimaging studies. Neurosci Biobehav Rev. 2012;36:2130–2142.

22. Rauch SL, Shin LM, Phelps EA. Neurocircuitry models of posttraumatic stress disorder and extinction: human neuroimaging research--past, present, and future. Biol Psychiatry. 2006;60:376–382.

23. Ressler KJ, Berretta S, Bolshakov VY, Rosso IM, Meloni EG, Rauch SL, et al. Post-traumatic stress disorder: clinical and translational neuroscience from cells to circuits. Nat Rev Neurol. 2022;18:273–288.

24. Shin LM, Rauch SL, Pitman RK. Amygdala, medial prefrontal cortex, and hippocampal function in PTSD. Ann N Y Acad Sci. 2006;1071:67–79.

25. Ressler KJ, Mercer KB, Bradley B, Jovanovic T, Mahan A, Kerley K, et al. Post-traumatic stress disorder is associated with PACAP and the PAC1 receptor. Nature. 2011;470:492– 497.

26. Stevens JS, Almli LM, Fani N, Gutman DA, Bradley B, Norrholm SD, et al. PACAP receptor gene polymorphism impacts fear responses in the amygdala and hippocampus. Proc Natl Acad Sci U S A. 2014;111:3158–3163.

27. Clancy KJ, Devignes Q, Kumar P, May V, Hammack SE, Akman E, et al. Circulating PACAP levels are associated with increased amygdala-default mode network resting-state connectivity in posttraumatic stress disorder. Neuropsychopharmacol. 2023;48:1245– 1254.

28. King SB, Toufexis DJ, Hammack SE. Pituitary adenylate cyclase activating polypeptide (PACAP), stress, and sex hormones. Stress. 2017;20:465–475.

29. Weathers FW, Bovin MJ, Lee DJ, Sloan DM, Schnurr PP, Kaloupek DG, et al. The Clinician-Administered PTSD Scale for DSM-5 (CAPS-5): Development and initial psychometric evaluation in military veterans. Psychol Assess. 2018;30:383–395.

30. Scrucca L, Fraley C, Murphy TB, Raftery AE. Model-Based Clustering, Classification, and Density Estimation Using mclust in R. New York: Chapman and Hall/CRC; 2023.

31. Esteban O, Markiewicz CJ, Blair RW, Moodie CA, Isik AI, Erramuzpe A, et al. fMRIPrep: a robust preprocessing pipeline for functional MRI. Nat Methods. 2019;16:111–116.

32. Pruim RHR, Mennes M, van Rooij D, Llera A, Buitelaar JK, Beckmann CF. ICA-AROMA: A robust ICA-based strategy for removing motion artifacts from fMRI data. Neuroimage. 2015;112:267–277.

33. Whitfield-Gabrieli S, Nieto-Castanon A. Conn: a functional connectivity toolbox for correlated and anticorrelated brain networks. Brain Connect. 2012;2:125–141.

34. Eickhoff S, Walters NB, Schleicher A, Kril J, Egan GF, Zilles K, et al. High-resolution MRI reflects myeloarchitecture and cytoarchitecture of human cerebral cortex. Hum Brain Mapp. 2005;24:206–215.

35. Amunts K, Kedo O, Kindler M, Pieperhoff P, Mohlberg H, Shah NJ, et al. Cytoarchitectonic mapping of the human amygdala, hippocampal region and entorhinal cortex: intersubject variability and probability maps. Anat Embryol (Berl). 2005;210:343–352.

36. Newcombe RG. Interval estimation for the difference between independent proportions: comparison of eleven methods. Statistics in Medicine. 1998;17:873–890.

37. Lenth RV. emmeans: Estimated Marginal Means, aka Least-Squares Means. 2017:1.11.2-8.

38. Eickhoff SB, Stephan KE, Mohlberg H, Grefkes C, Fink GR, Amunts K, et al. A new SPM toolbox for combining probabilistic cytoarchitectonic maps and functional imaging data. Neuroimage. 2005;25:1325–1335.

39. Margulies DS, Ghosh SS, Goulas A, Falkiewicz M, Huntenburg JM, Langs G, et al. Situating the default-mode network along a principal gradient of macroscale cortical organization. Proceedings of the National Academy of Sciences. 2016;113:12574–12579.

40. Salo T, Yarkoni T, Nichols TE, Poline J-B, Kent JD, Gorgolewski KJ, et al. neurostuff/NiMARE: 0.0.12rc7. 2022.

41. Salo T, Yarkoni T, Nichols T, Poline J-B, Bilgel M, Bottenhorn K, et al. NiMARE: Neuroimaging Meta-Analysis Research Environment. Aperture Neuro. 2023. 23 August 2023. 10.52294/001c.87480.

42. Yarkoni T, Poldrack RA, Nichols TE, Van Essen DC, Wager TD. Large-scale automated synthesis of human functional neuroimaging data. Nat Methods. 2011;8:665–670.

43. Ehlers A, Hackmann A, Steil R, Clohessy S, Wenninger K, Winter H. The nature of intrusive memories after trauma: the warning signal hypothesis. Behav Res Ther. 2002;40:995– 1002.

44. Adler LE, Pang K, Gerhardt G, Rose GM. Modulation of the gating of auditory evoked potentials by norepinephrine: Pharmacological evidence obtained using a selective neurotoxin. Biological Psychiatry. 1988;24:179–190.

45. Aston-Jones G, Rajkowski J, Kubiak P, Alexinsky T. Locus coeruleus neurons in monkey are selectively activated by attended cues in a vigilance task. J Neurosci. 1994;14:4467–4480.

46. Berridge CW, Waterhouse BD. The locus coeruleus–noradrenergic system: modulation of behavioral state and state-dependent cognitive processes. Brain Research Reviews. 2003;42:33–84.

47. Clancy K, Ding M, Bernat E, Schmidt NB, Li W. Restless ‘rest’: intrinsic sensory hyperactivity and disinhibition in post-traumatic stress disorder. Brain. 2017;140:2041– 2050.

48. Gelbard-Sagiv H, Magidov E, Sharon H, Hendler T, Nir Y. Noradrenaline Modulates Visual Perception and Late Visually Evoked Activity. Current Biology. 2018;28:2239–2249.e6.

49. McBurney-Lin J, Lu J, Zuo Y, Yang H. Locus coeruleus-norepinephrine modulation of sensory processing and perception: A focused review. Neuroscience & Biobehavioral Reviews. 2019;105:190–199.

50. Mehrpour V, Martinez-Trujillo JC, Treue S. Attention amplifies neural representations of changes in sensory input at the expense of perceptual accuracy. Nat Commun. 2020;11:2128.

51. Pessoa L, Kastner S, Ungerleider LG. Neuroimaging Studies of Attention: From Modulation of Sensory Processing to Top-Down Control. J Neurosci. 2003;23:3990–3998.

52. Southwick SM, Krystal JH, Bremner JD, Morgan CA III, Nicolaou AL, Nagy LM, et al. Noradrenergic and Serotonergic Function in Posttraumatic Stress Disorder. Archives of General Psychiatry. 1997;54:749–758.

53. Southwick SM, Bremner JD, Rasmusson A, Morgan CA, Arnsten A, Charney DS. Role of norepinephrine in the pathophysiology and treatment of posttraumatic stress disorder. Biological Psychiatry. 1999;46:1192–1204.

54. Bremner JD, Innis RB, Ng CK, Staib LH, Salomon RM, Bronen RA, et al. Positron Emission Tomography Measurement of Cerebral Metabolic Correlates of Yohimbine Administration in Combat-Related Posttraumatic Stress Disorder. Archives of General Psychiatry. 1997;54:246–254.

55. Kellner M, Levengood R, Yehuda R, Wiedemann K. Provocation of a posttraumatic flashback by cholecystokinin tetrapeptide? Am J Psychiatry. 1998;155:1299.

56. Nixon RDV, Bryant RA. Induced arousal and reexperiencing in acute stress disorder. Journal of Anxiety Disorders. 2005;19:587–594.

57. Stroth N, Holighaus Y, Ait-Ali D, Eiden LE. PACAP: a master regulator of neuroendocrine stress circuits and the cellular stress response. Ann N Y Acad Sci. 2011;1220:49–59.

58. Davis M. The role of the amygdala in fear and anxiety. Annu Rev Neurosci. 1992;15:353– 375.

59. Ago Y, Hayata-Takano A, Kawanai T, Yamauchi R, Takeuchi S, Cushman JD, et al. Impaired extinction of cued fear memory and abnormal dendritic morphology in the prelimbic and infralimbic cortices in VPAC2 receptor (*VIPR2*)-deficient mice. Neurobiology of Learning and Memory. 2017;145:222–231.

60. Cho J-H, Zushida K, Shumyatsky GP, Carlezon WA, Meloni EG, Bolshakov VY. Pituitary adenylate cyclase-activating polypeptide induces postsynaptically expressed potentiation in the intra-amygdala circuit. J Neurosci. 2012;32:14165–14177.

61. Meloni EG, Venkataraman A, Donahue RJ, Carlezon WA. Bi-directional effects of pituitary adenylate cyclase-activating polypeptide (PACAP) on fear-related behavior and c-Fos expression after fear conditioning in rats. Psychoneuroendocrinology. 2016;64:12–21.

62. Schmidt SD, Myskiw JC, Furini CRG, Schmidt BE, Cavalcante LE, Izquierdo I. PACAP modulates the consolidation and extinction of the contextual fear conditioning through NMDA receptors. Neurobiology of Learning and Memory. 2015;118:120–124.

63. Velasco ER, Florido A, Flores Á, Senabre E, Gomez-Gomez A, Torres A, et al. PACAP-PAC1R modulates fear extinction via the ventromedial hypothalamus. Nat Commun. 2022;13:4374.

64. Kirry AJ, Herbst MR, Poirier SE, Maskeri MM, Rothwell AC, Twining RC, et al. Pituitary adenylate cyclase-activating polypeptide (PACAP) signaling in the prefrontal cortex modulates cued fear learning, but not spatial working memory, in female rats. Neuropharmacology. 2018;133:145–154.

65. Rabellino D, Tursich M, Frewen PA, Daniels JK, Densmore M, Théberge J, et al. Intrinsic Connectivity Networks in post-traumatic stress disorder during sub- and supraliminal processing of threat-related stimuli. Acta Psychiatr Scand. 2015;132:365–378.

66. Ferri J, Schmidt J, Hajcak G, Canli T. Emotion regulation and amygdala-precuneus connectivity: Focusing on attentional deployment. Cogn Affect Behav Neurosci. 2016;16:991–1002.

67. Daselaar SM, Rice HJ, Greenberg DL, Cabeza R, LaBar KS, Rubin DC. The Spatiotemporal Dynamics of Autobiographical Memory: Neural Correlates of Recall, Emotional Intensity, and Reliving. Cerebral Cortex. 2008;18:217–229.

68. Mazzoni G, Clark A, De Bartolo A, Guerrini C, Nahouli Z, Duzzi D, et al. Brain activation in highly superior autobiographical memory: The role of the precuneus in the autobiographical memory retrieval network. Cortex. 2019;120:588–602.

69. Richter FR, Cooper RA, Bays PM, Simons JS. Distinct neural mechanisms underlie the success, precision, and vividness of episodic memory. eLife. 2016;5:e18260.

70. Young KD, Siegle GJ, Misaki M, Zotev V, Phillips R, Drevets WC, et al. Altered task-based and resting-state amygdala functional connectivity following real-time fMRI amygdala neurofeedback training in major depressive disorder. NeuroImage: Clinical. 2018;17:691– 703.

71. Burgos-Robles A, Vidal-Gonzalez I, Quirk GJ. Sustained Conditioned Responses in Prelimbic Prefrontal Neurons Are Correlated with Fear Expression and Extinction Failure. J Neurosci. 2009;29:8474–8482.

72. Etkin A, Egner T, Kalisch R. Emotional processing in anterior cingulate and medial prefrontal cortex. Trends in Cognitive Sciences. 2011;15:85–93.

73. Tovote P, Fadok JP, Lüthi A. Neuronal circuits for fear and anxiety. Nat Rev Neurosci. 2015;16:317–331.

74. Arruda-Carvalho M, Clem RL. Prefrontal-amygdala fear networks come into focus. Front Syst Neurosci. 2015;9.

75. Do-Monte FH, Quiñones-Laracuente K, Quirk GJ. A temporal shift in the circuits mediating retrieval of fear memory. Nature. 2015;519:460–463.

76. Cullen PK, Gilman TL, Winiecki P, Riccio DC, Jasnow AM. Activity of the anterior cingulate cortex and ventral hippocampus underlie increases in contextual fear generalization. Neurobiol Learn Mem. 2015;124:19–27.

77. de Lima MAX, Baldo MVC, Oliveira FA, Canteras NS. The anterior cingulate cortex and its role in controlling contextual fear memory to predatory threats. eLife. 2022;11:e67007.

78. Frankland PW, Bontempi B, Talton LE, Kaczmarek L, Silva AJ. The Involvement of the Anterior Cingulate Cortex in Remote Contextual Fear Memory. Science. 2004;304:881– 883.

79. Vetere G, Restivo L, Cole CJ, Ross PJ, Ammassari-Teule M, Josselyn SA, et al. Spine growth in the anterior cingulate cortex is necessary for the consolidation of contextual fear memory. Proceedings of the National Academy of Sciences. 2011;108:8456–8460.

80. Bone MB, Buchsbaum BR. Detailed Episodic Memory Depends on Concurrent Reactivation of Basic Visual Features within the Posterior Hippocampus and Early Visual Cortex. Cereb Cortex Commun. 2021;2:tgab045.

81. Clancy KJ, Devignes Q, Ren B, Pollmann Y, Nielsen SR, Howell K, et al. Spatiotemporal dynamics of hippocampal-cortical networks underlying the unique phenomenological properties of trauma-related intrusive memories. Mol Psychiatry. 2024;29:2161–2169.

82. Waldhauser GT, Braun V, Hanslmayr S. Episodic Memory Retrieval Functionally Relies on Very Rapid Reactivation of Sensory Information. J Neurosci. 2016;36:251–260.

83. Wheeler ME, Petersen SE, Buckner RL. Memory’s echo: Vivid remembering reactivates sensory-specific cortex. Proceedings of the National Academy of Sciences. 2000;97:11125–11129.

84. Kravitz DJ, Saleem KS, Baker CI, Mishkin M. A new neural framework for visuospatial processing. Nat Rev Neurosci. 2011;12:217–230.

85. Pearson J. The human imagination: the cognitive neuroscience of visual mental imagery. Nat Rev Neurosci. 2019;20:624–634.

86. Schubring D, Schupp HT. Emotion and Brain Oscillations: High Arousal is Associated with Decreases in Alpha- and Lower Beta-Band Power. Cerebral Cortex. 2021;31:1597–1608.

87. Breckel TPK, Giessing C, Thiel CM. Impact of brain networks involved in vigilance on processing irrelevant visual motion. NeuroImage. 2011;55:1754–1762.

88. Kastner S, De Weerd P, Desimone R, Ungerleider LG. Mechanisms of Directed Attention in the Human Extrastriate Cortex as Revealed by Functional MRI. Science. 1998;282:108– 111.

89. Akiki TJ, Averill CL, Abdallah CG. A Network-Based Neurobiological Model of PTSD: Evidence from Structural and Functional Neuroimaging Studies. Current Psychiatry Reports. 2017;19:81.

90. Akiki TJ, Averill CL, Wrocklage KM, Scott JC, Averill LA, Schweinsburg B, et al. Default mode network abnormalities in posttraumatic stress disorder: A novel network-restricted topology approach. Neuroimage. 2018;176:489–498.

91. Clancy KJ, Andrzejewski JA, Simon J, Ding M, Schmidt NB, Li W. Posttraumatic Stress Disorder Is Associated with α Dysrhythmia across the Visual Cortex and the Default Mode Network. eNeuro. 2020;7.

92. Miller DR, Hayes SM, Hayes JP, Spielberg JM, Lafleche G, Verfaellie M. Default Mode Network Subsystems Are Differentially Disrupted in Posttraumatic Stress Disorder. Biological Psychiatry: Cognitive Neuroscience and Neuroimaging. 2017;2:363–371.

93. Patel R, Spreng RN, Shin LM, Girard TA. Neurocircuitry models of posttraumatic stress disorder and beyond: A meta-analysis of functional neuroimaging studies. Neuroscience & Biobehavioral Reviews. 2012;36:2130–2142.

94. Pitman RK, Rasmusson AM, Koenen KC, Shin LM, Orr SP, Gilbertson MW, et al. Biological studies of post-traumatic stress disorder. Nat Rev Neurosci. 2012;13:769–787.

95. Sripada RK, King AP, Welsh RC, Garfinkel SN, Wang X, Sripada CS, et al. Neural dysregulation in posttraumatic stress disorder: evidence for disrupted equilibrium between salience and default mode brain networks. Psychosom Med. 2012;74:904–911.

96. Kearney BE, Terpou BA, Densmore M, Shaw SB, Théberge J, Jetly R, et al. How the body remembers: Examining the default mode and sensorimotor networks during moral injury autobiographical memory retrieval in PTSD. Neuroimage Clin. 2023;38:103426.

97. Kearney BE, Lanius RA. Why reliving is not remembering and the unique neurobiological representation of traumatic memory. Nat Mental Health. 2024;2:1142–1151.

98. Clancy KJ, Albizu A, Schmidt NB, Li W. Intrinsic sensory disinhibition contributes to intrusive re-experiencing in combat veterans. Sci Rep. 2020;10:936.

99. Brown VM, LaBar KS, Haswell CC, Gold AL, McCarthy G, Morey RA. Altered Resting-State Functional Connectivity of Basolateral and Centromedial Amygdala Complexes in Posttraumatic Stress Disorder. Neuropsychopharmacol. 2014;39:351–359.

100. Etkin A, Prater KE, Schatzberg AF, Menon V, Greicius MD. Disrupted Amygdalar Subregion Functional Connectivity and Evidence of a Compensatory Network in Generalized Anxiety Disorder. Archives of General Psychiatry. 2009;66:1361–1372.

101. Roy AK, Shehzad Z, Margulies DS, Kelly AMC, Uddin LQ, Gotimer K, et al. Functional connectivity of the human amygdala using resting state fMRI. NeuroImage. 2009;45:614– 626.

102. Roy AK, Fudge JL, Kelly C, Perry JSA, Daniele T, Carlisi C, et al. Intrinsic Functional Connectivity of Amygdala-Based Networks in Adolescent Generalized Anxiety Disorder. Journal of the American Academy of Child & Adolescent Psychiatry. 2013;52:290–299.e2.

103. Brewin CR. Re-experiencing traumatic events in PTSD: new avenues in research on intrusive memories and flashbacks. Eur J Psychotraumatol. 2015;6:10.3402/ejpt.v6.27180.

